# Effective coverage of diabetes and hypertension in Thailand: challenges and recommendations

**DOI:** 10.1101/2021.11.25.21266867

**Authors:** Nattadhanai Rajatanavin, Woranan Witthayapipopsakul, Vuthiphan Vongmongkol, Nithiwat Saengruang, Yaowaluk Wanwong, Aniqa Islam Marshall, Walaiporn Patcharanarumol, Viroj Tangcharoensathien

**Affiliations:** International Health Policy Program, Ministry of Public Health, Nonthaburi, Thailand; Global Health Divsion, Ministry of Public Health, Nonthaburi, Thailand

**Keywords:** Effective coverage, diabetes mellitus, hypertension, Thailand

## Abstract

**Introduction:** Increased Disability Adjusted Life Year (DALY) of diabetes and hypertension draws policy attention to improve effective coverage. This study assesses effective coverage of the two conditions in Thailand between 2016 and 2019.

**Method:** We estimated total diabetes and hypertension cases using age and sex specific prevalence rates for respective populations. Individual data from public insurance databases (2016-2019) were retrieved to estimate three indicators: detected need (diagnosed/total estimated cases), crude coverage (received health services/total estimated cases) and effective coverage (controlled/total estimated cases). Controlled diabetes was defined as Haemoglobin A1C (HbA1C) below 7% and controlled hypertension as blood pressure below 140/90 mmHg. In-depth interview of 85 multi-stakeholder key informants was conducted to identify challenges to better effective coverage.

**Results:** In 2016-2019, among Universal Coverage Scheme members residing outside Bangkok, estimated cases were around 3.1-3.2 million for diabetes and 8.7-9.2 million for hypertension. For diabetes services, all three indicators have shown slow increase over the four years (67.4%, 69.9%, 71.9%, and 74.7% for detected need; 38.7%, 43.1%, 45.1%, 49.8% for crude coverage; and 8.1%, 10.5%, 11.8%, 11.7% for effective coverage). For hypertension services, the performance was poorer for detection (48.9%, 50.3%, 51.8%, 53.3%) and crude coverage (22.3%, 24.7%, 26.5%, 29.2%) but was better for effective coverage (11.3%, 13.2%, 15.1%, 15.7%) than diabetes service. For both diseases, the estimates were higher for the females and older age groups than their counterparts. Complex interplays between supply and demand side barriers were a key challenge. Database challenges remain which hamper regular assessment of effective coverage.

**Conclusion:** Given the increased diabetes and hypertension prevalence, strategic recommendations cover long term actions for primary prevention of known risk factors as unhealthy diet and sedentary behaviour. Short term actions aim to improve effective coverage through the application of Chronic Care Model, increase attention to non-pharmacological intervention and patient empowerment.

**Summary Box:** *What is already known?:*

- As Effective coverage is the gains in health resulting from health interventions, the measure provides high granularity for assessing the performance of Universal Health Coverage.
- Non-Communicable Diseases (NCDs) account for more than two-thirds of global deaths, with Diabetes and hypertension as the two main drivers of NCD prevalence.
- The effective coverage of diabetes and hypertension are important measures of Thailand’s Universal Health Coverage performance.

*What are the new findings?:*

- The effective coverage of diabetes and hypertension in Thailand has shown slow increase over a period of four years, in every step of care. Interestingly, effective coverage among females and older populations are better than their counterparts.
- The key issues resulting in low effective coverage of both diabetes and hypertension are due to inconvenient hospital hours and high workload for health care workers leading to discontinuity of care, over-crowding and long waiting times; as well as patient unwillingness to modify behavioural aspects leading to NCD’s.
- The major barriers to measuring effective coverage in Thailand is the lack of valid, complete, and accurate data.

*What do the new findings imply?:*

- Despite 17 years of Universal Health Coverage, NCDs care in Thailand is still underperformed with slow improvement year by year.
- Policies to address the effective coverage of diabetes and hypertension should 1. Aim to increase patient awareness and screening of NCD’s; 2. Focus on non-pharmacological intervention and patient empowerment; 3. Utilize digital technology and innovations to support the current over-stretched health personnel; 4. Strengthen campaigns to modify patient lifestyles; 5. Initiate and support population-based strategies addressing NCD risk factors.

## 1. INTRODUCTION

Effective Coverage, defined as the “fraction of real health gained from an intervention”, can be used as an indicator to measure health system performance [1,2]. In contrast to traditional measures of crude coverage which simply captures access to health services among those who need health interventions, effective coverage takes into account health gain as a result of those health interventions. Effective coverage can be applied to measure the overall performance of a health system or selected conditions based on priority burden of diseases.

As countries advance towards Universal Health Coverage (UHC), moving from measuring conventional crude coverage of health services towards effective coverage has been emphasised [3,4]. Effective coverage provides high granularity for assessing UHC performance [2].

Thailand implemented UHC in 2002, achieving full population coverage through three publicly financed insurance schemes. Civil Servant Medical Benefit Scheme (CSMBS) covers around 4.4 million of civil servants and their dependants managed by Comptroller General’s Department; Social Health Insurance (SHI) covers 10.6 million of private sector employees managed by the Social Security Office; and the Universal Coverage Scheme (UCS) is for the remaining 48 million population, managed by the National Health Security Office [5]. However, whether the population has achieved better health outcomes as a result of UHC is unknown, therefore warranting studies on the effective coverage of health services under UHC.

Non-Communicable Diseases (NCDs) account for more than two-thirds of global deaths, with a rising burden of disease [6]. Diabetes and hypertension are the two main drivers of NCD prevalence globally, with an estimated 463 million adults living with diabetes in 2019 and 1,130 million adults living with hypertension in 2015 [7,8]. In Thailand, according to the National Health Examination Survey (NHES) the prevalence of diabetes among Thai adults has increased from 6.8% in 2004 to 8.9% in 2014, while hypertension has increased from 21.4% to 24.7% in the same period [9]. Diabetes ranks first among females and seventh among males for Disability Adjusted Life Year (DALY) loss according to the Burden of Disease report in 2014 [10,11]. Given the significant burden of disease, maximizing effective coverage of diabetes and hypertension is a key policy priority for the Thai Government. Both diabetes and hypertension are among national priority diseases according to the twelfth five-year National Health Development Plan (2017-2021) [12].

In assessing effective coverage, three important indicators must be defined and investigated: 1. detected need, 2. crude coverage and 3. effective coverage. Due to the variations in available data and the desirable health outcomes across different effective coverage studies, there is no universal criterion for each indicator [1]. For most studies on diabetes, total estimated diabetes cases were from survey results or calculated from diabetic prevalence rates in the area, which was defined as patients diagnosed by a doctor, taking medication, or having fasting plasma glucose (FBG), oral glucose tolerance test or Haemoglobin A1C (HbA1C) test above a defined cut-off point [13–18]. Therefore, detected need has been defined as proportion of diagnosed diabetes patients by the total estimated diabetes cases. [13–18]. Crude coverage of diabetes is generally defined as the proportion of diabetic patients receiving medication or lifestyle modification interventions by the total estimated diabetes cases. [13–18]. Effective coverage for diabetes has been defined as the proportion of treated patients having achieved desirable cut-off points for FBG or HbA1C by the total estimated diabetes cases [13–18]. In another study conducted in Mexico, researchers subcategorized effective coverage into multiple indicators including blood sugar control, lipid control as well as other diabetic complications [19].

Studies on hypertension also utilized total estimated hypertension cases from survey results or calculated from hypertension prevalence rates in the area, which defined patients as those diagnosed by doctors, prescribed with anti-hypertensive medication, or having blood pressure of greater than or equal to 140/90 mmHg, [16,20–23]. Detected need has been defined as the proportion of diagnosed hypertensive patients by the total estimated hypertensive cases. [16,20–23]. In some studies, crude coverage is defined as the proportion of patients prescribed with anti-hypertensive medication or life-style modifications by the total estimated hypertensive cases; while effective coverage is defined as the proportion of hypertensive patients who received treatment with a blood pressure less than 140/90 mmHg by the total estimated hypertensive cases.[16,20–23].

The data obtained for the calculation of effective coverage highly varies across different studies. In countries such as the Republic of Korea, Chile, and Mexico, individual level health service database is commonly utilized, while other countries conduct health surveys to estimate health status in the population [16,18–22].

This study aims to estimate effective coverage indicators for diabetes and hypertension using administrative health service databases in Thailand. In addition, we seek to understand the barriers to greater level of effective coverage and the challenges of regular assessment of effective coverage as one measure to monitor health system performance using multistakeholder semi-structured interviews.

## 2. METHODS

A mixed method research design was applied using secondary data analysis of administrative health service datasets and key informant interviews. Ethics approval was obtained from the Institute for the Development of Human Research Protections (IHRP) reference IHRP2019030, obtained on the 27^th^ of March 2019.

### 2.1 Secondary data analysis

This quantitative analysis aims to estimate effective coverage indicators for diabetes and hypertension, disaggregate by sex and age group, and describe the time trend.

#### Study populations and data sources

Our study populations were diabetes and hypertension patients aged 15 and older who were members of the Universal Coverage Scheme outside of Bangkok between 2016 and 2019. We excluded those registered with health facilities located in Bangkok because of limitation in accessing the data and populations aged less than 15 years old because there was no prevalence data for children.

Four consecutive years, 2016-2019, of three sets of databases were retrieved and analysed. 1) Out-patient health prevention and promotion database held by the National Health Security Office with individual patient health records for each out-patient visit. 2) Electronic claim database held by the National Health Security Office with claim data from healthcare facilities for inpatient and high-cost outpatient health services. 3) Civil registration database held by the Ministry of Interior with number of populations by age and sex for estimating total diabetes and hypertension cases in the population.

#### Defining effective coverage indicators

In this study, the three indicators (detected need, crude coverage and effective coverage) for our measurement were guided by previous literature, taking into account the data availability in Thailand, and were peered by relevant clinical specialists and Ministry of Public Health officers [9,24–29]. There are four sets of aggregated data required to estimate the three indicators.

**Estimated cases** are the number of total national diabetes and hypertension individuals estimated using the age and sex specific prevalence rates from the NHES 2014 (the latest available at the time of study) multiplied by the number of respective UCS populations in 2016-2019. Diabetes cases in the NHES 2014 were defined as patients diagnosed by doctors, taking medication, or with a FPG ≥ 126 mg/dL, while hypertension cases were defined as patients diagnosed by doctors, taking medication, or with a blood pressure ≥ 140/90 mm/Hg [9]. **Diagnosed cases** are the number of patients diagnosed with diabetes or hypertension based on the International Classification of Diseases (ICD10), “E10-E14” for diabetes and ICD10 “I10-I14” for hypertension. **Patients who received health services** are diagnosed cases who visited health facilities at least four times in a year. We used the four visits based on the average 3-month interval of diabetes and hypertension health service visits in Thailand [29]. **Controlled cases** are those who received health services and had the most recent HbA1C level of less than 7% for diabetes or the latest two consecutive visit blood pressure value of less than 140/90 mmHg for hypertension, which are benchmarks produced by Ministry of Public Health (MOPH) Key Performance Indicators and Thailand Clinical Practice Guidelines [24–26].

We applied the following definition and formula. **Detected need** is the proportion of diagnosed cases in the total estimated cases; **crude coverage** is the proportion of the patients who received health services in the total estimated cases; and **effective coverage** is the proportion of controlled cases in the total estimated cases. The operational definitions of the numerators and denominator for all indicators are summarised in Table 1. Stata 14.0 (StataCorpLP,TX Serial number: 401406358220) was used for data analysis.

**Table 1.** Definition and formulae of three key indicators for diabetes and hypertension used by this study

|  | Detected Need | Crude Coverage |
| --- | --- | --- |
| **Definition** | Proportion of diagnosed cases to total national estimated cases | Proportion of patients who have received continued health services to the total national estimated cases |
| **Diabetes** | $\frac{\# \text{ Patients Diagnosed with ICD 10 E10 – E14}}{\# \text{ Estimated Cases of Diabetes}}$ | $\frac{\# \text{ Patients that recieved health services } \geq 4 \text{ times per year}}{\# \text{ Estimated Cases of Diabetes}}$ |
| **Hypertension** | $\frac{\# \text{ Patients Diagnosed with ICD 10 I10 – I14}}{\# \text{ Estimated Cases of Hypertension}}$ | $\frac{\# \text{ Patients that recieved health services } \geq 4 \text{ times per year}}{\# \text{ Estimated Cases of Hypertension}}$ |
| **Effective Coverage** |  |  |
| **Definition** | Proportion of patients who have received continuous health service and demonstrated health gains to the total national estimated cases |  |
| **Diabetes** | $\frac{\text{Patients that recieved health services } \geq 4 \text{ times per year AND have a latest HbA1C result of } < 7\%}{\# \text{ Estimated Cases of Diabetes}}$ | |
| **Hypertension** | $\frac{\text{Patients that recieved health services } \geq 4 \text{ times per year AND have a BP result of 140/90 in the last 2 visits}}{\# \text{ Estimated Cases of Hypertension}}$ | |

### 2.2 Key Informant Interviews

Semi-structured key informant interviews were conducted between May and August 2019. The aims of the interviews were to discuss two key topics: barriers to greater level of effective coverage for diabetes and hypertension service in Thailand and challenges in measuring effective coverage.

A total of 85 key informants from four groups of experts representing Central and Regional health areas participated in this study to provide diverse and relevant information applicable to Thailand as a whole. Clinical specialists were identified from a list of university professors with expertise in managing diabetes and/or hypertension and experts from the National Diabetic Society and National Hypertension Society. Healthcare providers and public health officers were selected from the provinces with high, moderate and low performance in managing diabetes and hypertension against the MOPH Key Performance Indicators (KPI) on diabetes and hypertension control in 2018 [26]. Academics and experts on public health insurance were purposively selected from the MOPH’s technical departments and each of the three public health insurance agencies, respectively. See S1 Table.

Written informed consent and permission to audio record were sought prior to each interview. Interview records were transcribed. Two researchers extracted the transcribed data through a qualitative thematic analysis approach and final analysis was conducted through discussion among researchers.

## 3. Results

The Universal Coverage Scheme excluding Bangkok comprised of approximately 34.1-34.5 million adult populations during 2016-2019, with a balanced male-female ratio. Across the four years, nearly 80% were aged between 15-60 years and around 21-24% were elderly aged 60 and over. See Table 2

**Table 2.** Characteristics of the study population who are UCS members residing outside Bangkok, 2016-2019, and prevalence of Diabetes and Hypertension by Age & Sex

| Characteristics/Fiscal year (FY) |  |  | 2016 (n, %) |  | 2017 (n, %) |  | 2018 (n, %) |  | 2019 (n, %) |  |  |  |  |
| --- | --- | --- | --- | --- | --- | --- | --- | --- | --- | --- | --- | --- | --- |
| Total |  |  | 34,189,536 |  | 34,218,161 |  | 34,538,221 |  | 34,112,609 |  |  |  |  |
| Sex |  |  |  |  |  |  |  |  |  |  |  |  |  |
| Female |  |  | 17,322,452 (51%) |  | 17,332,181 (51%) |  | 17,323,392 (50%) |  | 17,249,648 (51%) |  |  |  |  |
| Male |  |  | 16,867,084 (49%) |  | 16,885,980 (49%) |  | 17,214,829 (50%) |  | 16,862,961 (49%) |  |  |  |  |
| Age group |  |  |  |  |  |  |  |  |  |  |  |  |  |
| 15 to less than 60 years |  |  | 26,932,819 (79%) |  | 26,692,965 (78%) |  | 26,717,187 (77%) |  | 25,972,544 (76%) |  |  |  |  |
| 60 years and older |  |  | 7,256,717 (21%) |  | 7,525,196 (22%) |  | 7,821,034 (23%) |  | 8,140,065 (24%) |  |  |  |  |
| Prevalence |  |  |  |  |  |  |  |  |  |  |  |  |  |
| Age group |  | 15-29 |  | 30-44 |  | 45-59 |  | 60-69 |  | 70-79 |  | ≥ 80 |  |
| Sex |  | M | F | M | F | M | F | M | F | M | F | M | F |
| Diabetes |  | 1.50% | 2.80% | 5.00% | 5.60% | 11.80% | 12.40% | 16.10% | 21.90% | 15.00% | 21.80% | 12.40% | 11.30% |
| Hypertension |  | 5.90% | 2.00% | 22.00% | 11.70% | 33.40% | 32.10% | 47.20% | 49.50% | 52.60% | 60.10% | 58.70% | 68.90% |

### 3.1 Total diabetes and hypertension cases among the UCS members

Of total UCS members residing outside of Bangkok, the total estimated cases were 3.2 million for diabetes and 9.2 million for hypertension in 2019. Estimated cases of diabetes and hypertension in 2019-2019, stratified by age and sex is presented in S2 and S3 Table.

### 3.2 Effective coverage estimates

In 2019, of the total 3.2 million estimated diabetes cases, 74.7% had been diagnosed and 49.8% received continued health services of at least 4 visits per year. Of the 9.2 million total estimated cases of hypertension; 53.3% were diagnosed and 29.2% received treatment during at least four health visits per year. See Fig 1.

**Figure 1.**
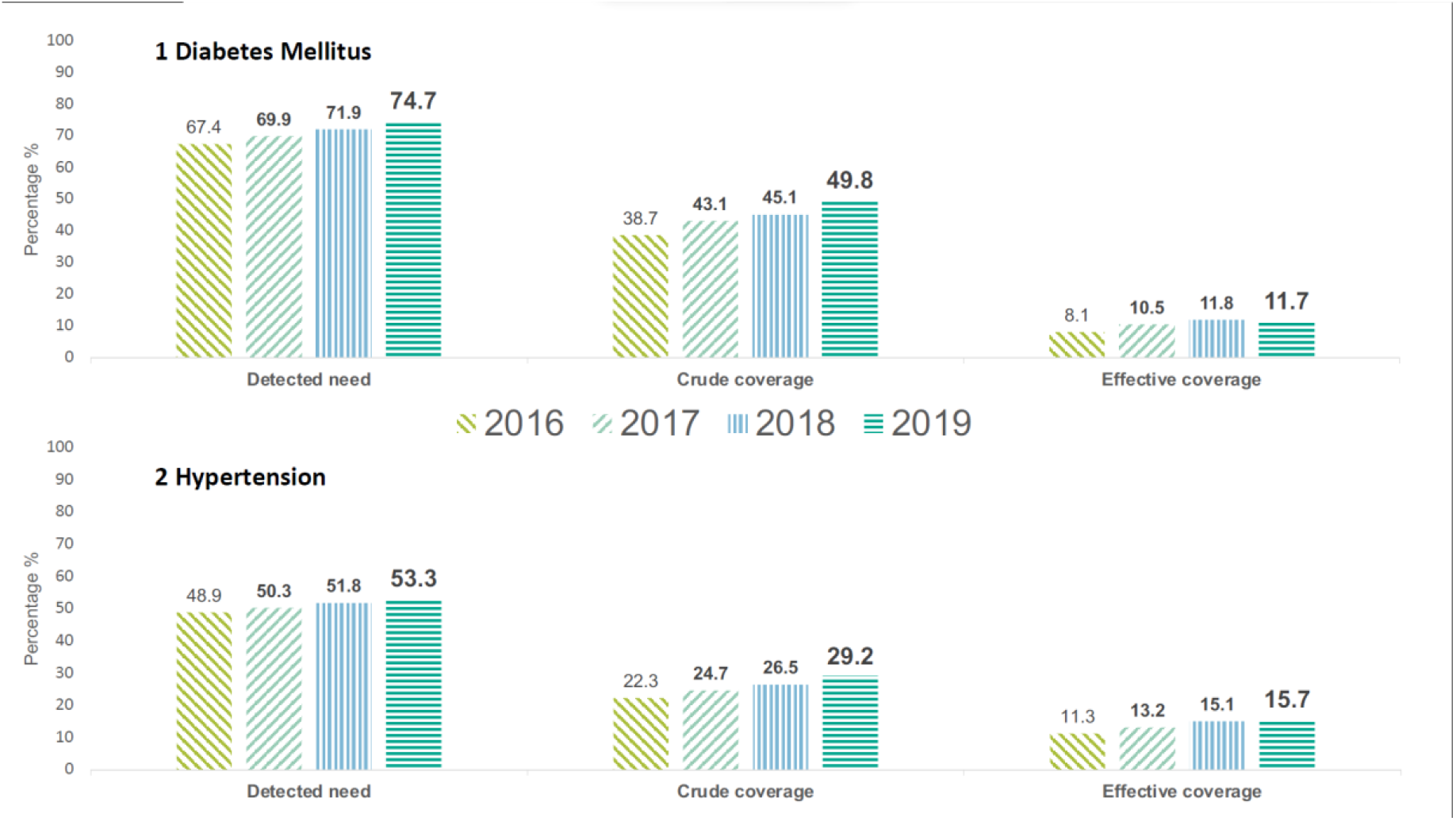
Effective coverage indicators for 1. diabetes and 2. hypertension 2016-2019

Based on these values, the effective coverage was estimated as 11.7% of total estimated cases with a HbA1C benchmark of <7% for diabetes, and 15.7% of total estimated cases for hypertension according to the benchmark of least four visits and controlled blood pressure below 140/90 mmHg in the last two consecutive visits.

### 3.3 Four-Year trend analysis

A four-year increasing trend of all indicators between 2016 and 2019 was found for both diabetes and hypertension.

Diabetes detected need increased by 7.3 percentage points from 67.4% to 74.7% while the crude coverage increased at a faster pace, by 11.1 percentage points from 38.7% to 49.8%. The effective coverage of diabetes increased at a much slower pace; 3.6 percentage points, from 8.1% to 11.7%. Fig 1 illustrates a large gap between crude and effective coverage; up to 38.1 percentage points in 2019.

The detected need of hypertension increased by 4.4 percentage points from 48.9% to 53.3%, the crude coverage increased at a slight faster pace, by 6.9 percentage points from 22.3% to 29.2%, while the effective coverage increased by 4.4 percentage points, from 11.3% to 15.7%. In 2019, there was a larger gap between detected need and crude coverage of 24.1 percentage points compared to the gap between crude and effective coverage at 13.5 percentage points.

### 3.4 Age-Sex stratification

Older age group, 60 years and older, was found to have higher detected need rate for both diabetes and hypertension. In 2019, diabetes rates were higher by 39.7 percentage points for detected need,

30.4 percentage points for crude coverage and 11.1 percentage points for effective coverage compared to those aged under 60 years. See Fig 2. Hypertension rates were higher by 35.3 percentage points for detected need, 22.4 percentage points for crude coverage and 11.5 percentage points for effective coverage compared to those aged less than 60 years. See Fig 3. Note that populations younger than 60 years consist of 54.6% of total estimated diabetes cases in the country (1.7 out of 3.2 million) and 52.6% of total estimated hypertension cases in the country (4.8 out of 9.2 million). See S2 and S3 Table.

**Figure 2.**
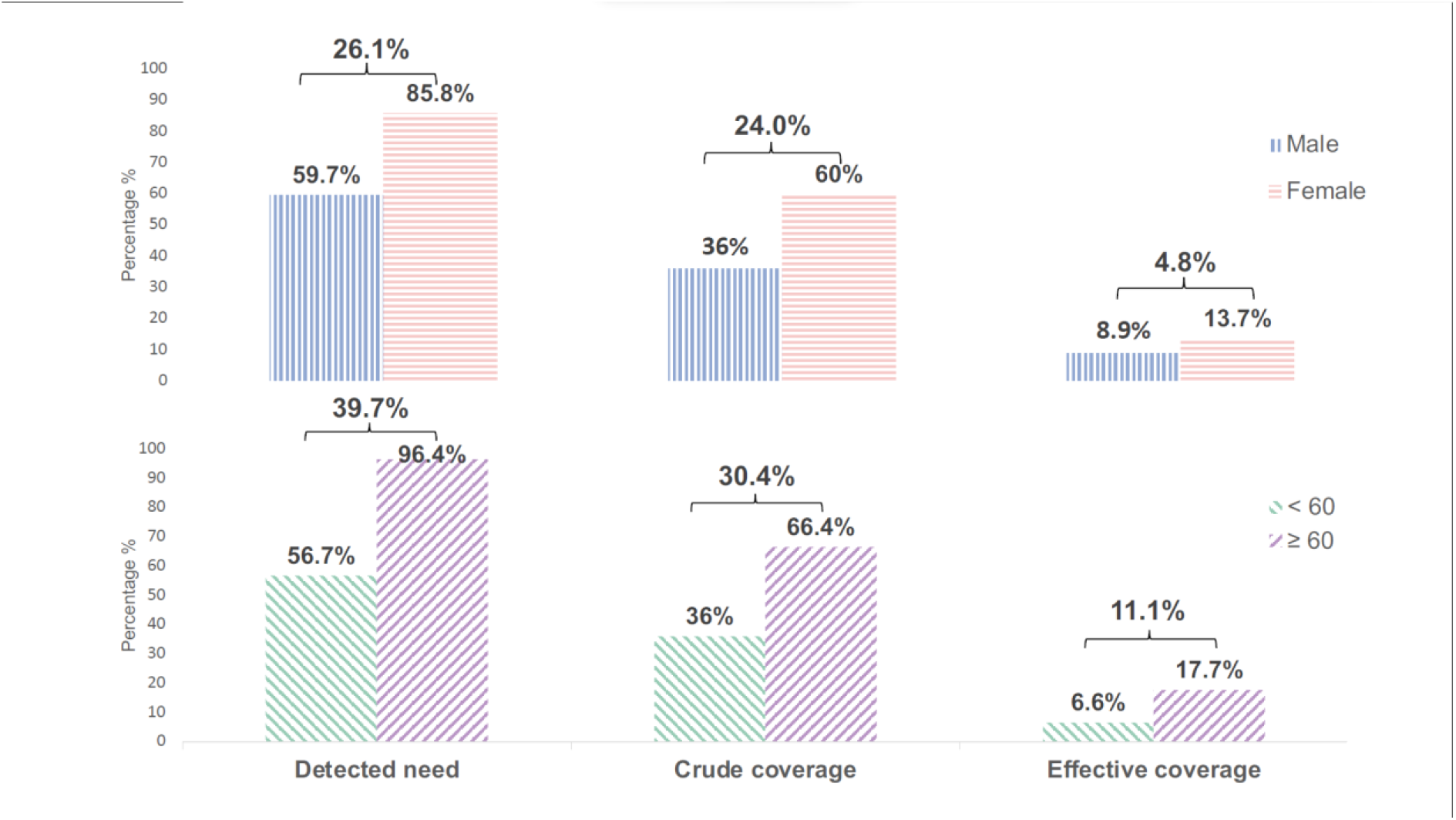
Effective coverage indicators for diabetes stratified by sex and age group

**Figure 3.**
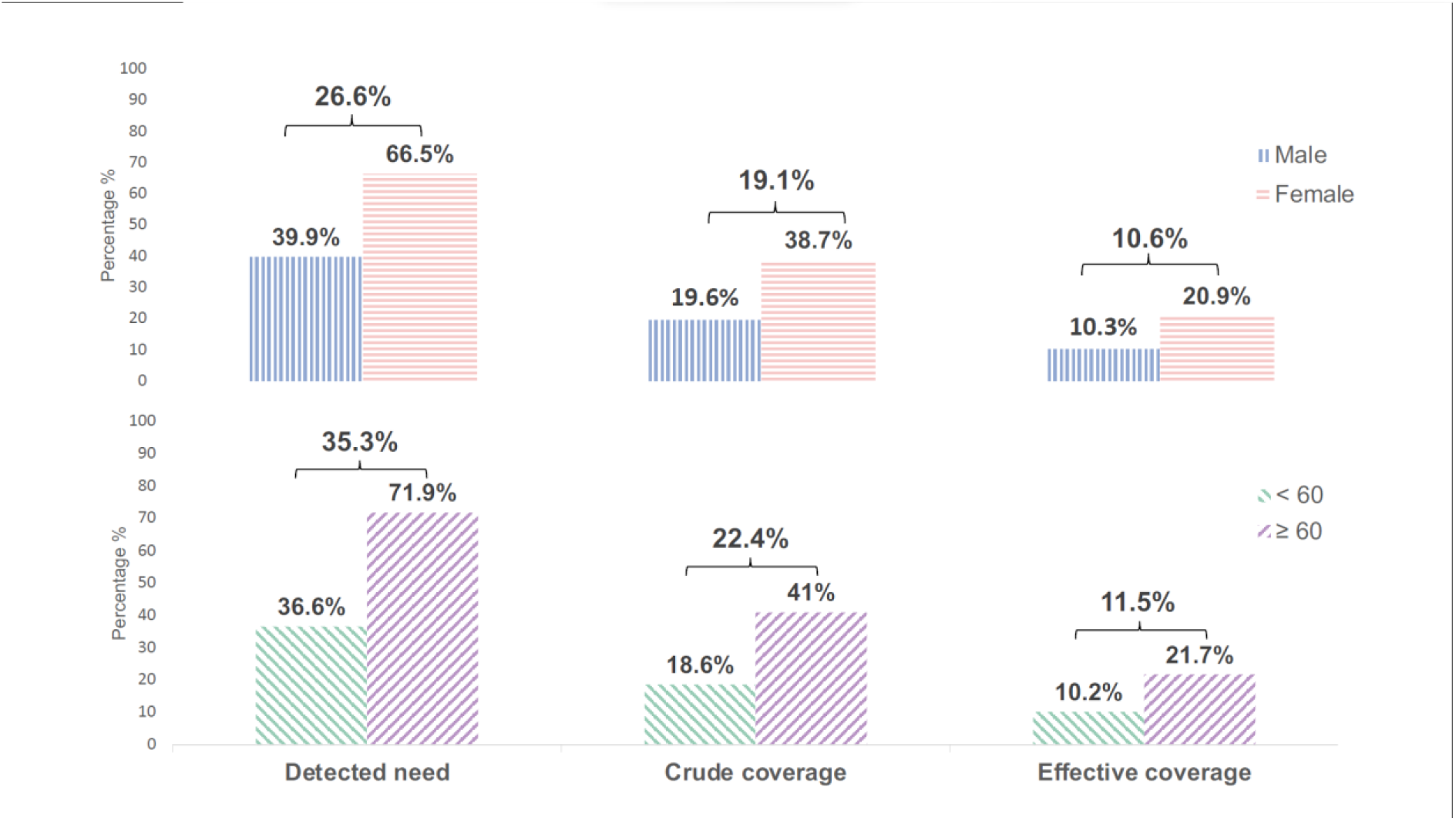
Effective coverage indicators for hypertension stratified by sex and age group

Females outperformed males in all three coverage indicators for both diabetes and hypertension. Diabetes rates were higher by 26.1 percentage points for detected need, 24 percentage points for crude coverage, and 4.8 percentage points for effective coverage in 2019. See Fig 2. Hypertension rates were higher by 26.6 percentage points for detected need, 19.1 percentage points for crude coverage, and 10.6 percentage points for effective coverage See Fig 3.

Age-Sex analysis in 2016, 2017 and 2018 yielded similar results. See S2 and S3 Table.

### 3.5 Health systems: barriers to effective coverage

In-depth interviews with key informants identified several health systems bottlenecks to achieve high level effective coverage for both conditions.

NCDs clinics in public health facilities, operate 8-16 hours on weekdays, is inconvenient for patients who are still working especially under 60 years; this leads to a loss to follow up and discontinuity of treatment. Community screening by sub-district health centres for adults > 35 years, one of the main approaches to identify new cases cannot cover all target populations, especially among the working age and those live-in urban areas. UCS implements capitation contract model for outpatient services, which requires UCS members to register with a primary healthcare network-consisting of health centres and a district hospital in rural area, or health centres and provincial hospitals in urban areas), where all services including NCD screening need to seek from this network.

To facilitate internal migration by the working age who seek job in urban cities outside their domicile districts, re-registration with a new network in the city enables access to care by these members, however, the re-registration process is not convenient for the patient. Despite these efforts, effective coverage among younger age group is low. Further, NCD is among all competing health agendas from the healthcare provider perspectives. Significant increased health service workloads result in over-crowding, long waiting times and inadequate times for quality counselling in particular non-pharmacological interventions to NCDs patients

> *“Urban settings are different. Some communities shut the door on us when we went out for screening diabetes and hypertension, some even threatened to take legal actions against trespassing. Even though we are health officers, if they do not welcome us, we cannot do anything, especially NCDs in condominiums.” [KI216 District Health Office]*
>
> *“For those who have been diagnosed, treated, but loss to follow-up, we do not have the ability to track them. Some work outside the community” [KI218 Provincial Health Office]*

On the demand-side, lifestyle and patient behaviour also limits effective coverage. Some patients do not cooperate to follow up or adhere to lifestyle modification, such as continue to smoke and drink alcohol, while providers do not have enough time for counselling

> *“Health education cannot change their lifestyle. Patient knows everything but does not follow our recommendations.” [KI240 District Health Office]*

Although a sub-district health promotion fund, jointly invested by the National Health Security Office and local government has been allocated for specific interventions relevant to local context; staff at sub-district health centers have yet to maximize use of this fund to counteract the social determinants of NCDs such as tobacco, alcohol, and unhealthy diet.

> *“The local health promotion fund aims to empower the local community to solve priority health problems. It has yet to overcome bureaucratic rigidity in using this fund” [KI405 Health Insurance]*

### 3.6 Information systems: barriers to measuring effective coverage

#### 3.5.1 Health data system

Staff at sub-district health centres and district hospitals are responsible for entering all data in the electronic recording system for NCDs such as detection, registration, visits for treatment and outcomes in term of HbA1c and blood pressure linking each record to each patient unique citizen identification number. However, limited human capacity, inadequate skill on information technology, lack of support and supervision result in inadequate validity and completeness of data systems for performance assessments.

> *“We try to learn, however, the process of data entry changes quite often. There are massive details which we need to learn by ourselves. Provincial or District IT experts never comes to help.” [KI213 Health Centre]*
>
> *“We do not have enough manpower at the health centres. We hire contractor workers for data entry. Some of them are high school graduated unemployed relatives of VHV” [KI223 Health Centre]*

#### 3.5.2 Governing the health systems

The MOPH set high KPI standards without thorough consultation with local health workers to probe its feasibility and challenges. From 2016 to 2019, screening target for diabetes and hypertension was set at > 90% of the population aged 35 and over, while the control target was set at ≥ 40% for diabetes (with HbA1C < 7%) and ≥ 50% for hypertension (with blood pressure <140/90 mmHg) [26]. This puts significant pressure on local health officers to report untrue higher data especially in the context of inadequate audit capacity.

> *“MOPH sets too high KPIs, sometimes not realistic, MPOH should consult with the local. Otherwise, if the KPI is unrealistically high, the MOPH may receive the high-performance data, but reliability remains a question.” [KI216 District Health Office]*

Additionally, there are no KPIs for continuous treatment rate or loss to follow up rates, which are crucial for performance assessments. The current fragmentation of information systems is complicated by inadequate Health Information Exchange system among health facilities to provide seamless care.

## 4. Discussion

### 4.1 Detected need, crude coverage and effective coverage

Though this study demonstrates an increasing trend of all three indicators (detected needs, crude coverage, and effective coverage) between 2016 and 2019; the level of effective coverage is still low, 11.7% for diabetes and 15.7% for hypertension in 2019. Diabetes service shows lower effective coverage than hypertension service in Thailand which is similar to the situations in the Republic of Korea but in contrast with those in South Africa where diabetes service showed better coverage performance. However, studies from different countries did not apply the same methodology [14,17,22,23].

Effective coverage estimates from this study is lower than the results reported by the NHES 2014, which found detected need, crude coverage, and effective coverage for diabetes 56.9%, 54.1%, and 23.5% respectively. For hypertension, the detected need, crude coverage, and effective coverage results from the NHES 2014 were 55.3%, 49.2%, and 29.7%, respectively. Similar to our findings, the NHES 2014 also reports higher effective coverage for those 60 years and older and females compare to their counterparts for both diabetes and hypertension. However, the definition for each indicator applied by the NHES 2014 is different from this study. In the NHES 2014, total estimated cases were calculated from actual number of patients in the survey. Detected need was defined as the proportion of patients previously diagnosed by a doctor or taking medication to the total estimated cases; crude coverage was defined as the proportion of patients who received medication to the total estimated cases; and effective coverage was defined as the proportion of patients who received medication and had a FBG < 130 mg/dL for diabetes and BP < 140/90 mm/Hg for hypertension in the survey to the total estimated cases. Effective coverage results is higher than results from our study due to less stringent definitions applied for the indicators [9].

A study in Thailand in 2018 using samples from hypertension screening programs and only recruiting undiagnosed population reports 49.9% effective coverage[30]. This study cannot be compared with our results due to different study populations.

The current community-based NCD screening is provided through health care workers in more than 8,000 sub-district health centres with support by village health volunteers, is not convenient for working populations [31]. The process involves community screening then confirmation by a doctor in district hospitals in 1-2 weeks. This long lead time results in loss to follow up. There is no screening service available in hospitals, clinics, pharmacies for walk-in and after hour services. These result in significant lower detected need in working age than older age group.

Even though continuous care is the key to well controlled diabetes and hypertension, many patients are lost to follow-up [24,25,27,28]. There is a similar large gap between detected need and crude coverage in 2019, 24.9 percentage points for diabetes and 24.1 percentage points for hypertension. See Fig 1. Such gaps are caused by demand and supply side barriers. On the patient side, regular visits, though four times a year, can be a burden from taking time off work among working age patients. Moreover, patients are complacent as there is no significant symptom or unconvinced in the benefits of treatment [32,33]. On the provider side, diabetes and hypertension rank first and second highest workload in outpatient departments, accounting for around 30% of outpatient visits to MOPH hospitals which equal to 79 million visits a year in 2019 [34].

In order to improve quality of care for chronic disease patients, a multidisciplinary team and Chronic Care Model--a collaborative partnership among patients, providers and health system, which incorporates a multilevel approach are required particularly for diabetes patients whose behaviours plays an important role in treatment outcomes [24,25,27,28,35–37]. However, the heavy work-load for hospitals prevents health care workers from spending adequate time for counselling on non-pharmacologic interventions. Further, there are limited choices of diabetes drugs (15 medicines) in the National List of Essential Medicine compared with hypertension (25 medicines) [38]. This results in large gap between crude and effective coverage in 2019, 38.1 percentage points for diabetes and 13.5 percentage points for hypertension. See Fig 1.

Patient’s attitude and perception play significant role. A qualitative study in Thailand found that patients’ experiences of severe complications from diabetes have better medication adherence, while lacking overt symptoms results in complacency, and that “normal” blood sugar levels are interpreted by patients as cure and there is no needs for continued medication [39]. Adequate diabetic health literacy and better glycaemic control are highly correlated [40]. A study from Thailand showed 61% of 312 type 2 diabetes patients had poor glycaemic control, as two-thirds of them had moderate health literacy levels [41]. Another study showed 48.7% of Thai hypertensive patients had inadequate health literacy [42].

Though inconvenient service hours can explain the low coverage indicators among working-age patients; underlying factors on gender disparity is unknown, warranting further investigation such as gender specific awareness and health literacy.

The concern of increased prevalence of diabetes (from 6.8% in 2004 to 8.9% in 2014) and hypertension (from 21.4% to 24.7% in the same period) is complicated by the low effective coverage of both conditions. Primary prevention is a key policy choice. Cochrane review reports no firm evidence that diet or physical activity alone can influence the risk of Type 2 diabetes mellitus (T2DM); but diet plus physical activity reduces or delays the incidence of T2DM in people with impaired glucose tolerance [43]. Healthy diet and physical activity promotions are active campaign against NCD supported by Thai Health Foundation established since 2001 [44]. Thailand may capitalize the high prevalence of adequate moderate-to-vigorous physical activities among Thai adults; it increased from 66.6% in 2012 to 70.1%, 69.5 %, 73.1%, 70.6%, 73.0%, 75.6% and 74.3% in 2013–2019 [45]. Additionally, Thailand has implemented many WHO recommended NCDs measures such as introducing sugar sweetened beverages tax, strengthening food environment, regulation of food marketing, alcohol and tobacco control [46–48]. However, significant efforts are required to reverse the worsening trend of diabetes and hypertension in Thailand.

### 4.2 Methodological challenges

This study uncovers methodological challenges. There is no consensus on how effective coverage indicators of diabetes and hypertension services are define [1]. There are many options and cut-off point that researcher may use. Our study defines crude coverage as those who visited health facilities at least four times in a year. For effective coverage, we rely on two surrogate outcomes with cut points. However, other choices of indicators such as ‘on medication’, ‘ever visit a doctor’ for crude coverage or ‘fasting blood sugar level below 126 mg/dL’, ‘no complications’, ‘HbA1C < 6.5%’ are also conceptually and clinically acceptable.

Feedback from peer review process suggested that selection of any surrogate parameters must consider not only clinical guidelines but availability of data. It should also be acknowledged that using surrogate outcomes, despite being easily measurable, may not always lead to individual health gain as wished by effective coverage concept.

A lack of standardized methodology leads to different studies employing varying definition and methods which makes cross study comparison, systematic review, or meta-analysis problematic. We recommend that a country aiming to use effective coverage for policy decision must seek for a methodology consensus among stakeholders prior to data collection. Factors to be considered include choice of indicators and cut-off points, data feasibility, interpretation capacity, and acceptance by stakeholders. A standardized methodology, at least at country level, will allow performance evaluation across time periods comparable and facilitate performance improvement.

### 4.3 Limitations

Key limitations relate to database and methodology. First, this study does not represent Thailand as it covers only patients under UCS. Two other schemes CSMBS and SHI do not have such records. This study also excludes 6 million population in Bangkok due to lack of data.

Second, there is no rigorous assessment of the validity of records such as ICD 10 diagnosis or biological markers. The inter-connectivity of database is a key gap for maximize use by healthcare providers in clinical service provision and monitoring outcome. High level of key performance indicators can lead to data creeping as data audits are inadequate.

Third, the national health examination survey does not cover population under 15 years old. Its five-year survey interval cannot produce prevalence rate applied as denominators for all three indicators in a timely manner.

Fourth, even though using HbA1C at <7% at the last visit is simple and easy to understand by policy makers, it is not in line with the current clinical practice which recommends 3-6 months regular monitoring of HbA1C according to biologic and socioeconomic profile of each patient. For hypertension, the well accepted blood pressure target of below 140/90 mmHg requires more points of blood pressure monitoring.

Finally, incompleteness of database was identified in both diseases. In order to calculate crude coverage, the total number of health facilities visits per year is required, however, data in 2016-2019 revealed only 84.1-90.4% and 75.6-84.0% of diagnosed diabetes and hypertension patients had complete health facilities visits data. In the same period, 28.6-41.1% of diabetes patients and 9.1-16.8% of hypertension patients, who received health services (having ≥ 4 times health facilities visits per year) did not have their HbA1C or blood pressure records, which limited the ability to calculate effective coverage estimates. As a result, this undetermined whether patients missed health service visits or whether visits were not recorded. Although the problem is gradually improving over the years, the incompleteness of data hampers accurate estimates of effective coverage.

## 5. Conclusion and recommendations

Our study found that effective coverage estimates for diabetes and hypertension service in Thailand were low despite showing an increasing trend. Three indicators, detected need, crude coverage and effective coverage, suggests large gaps along service cascades which are influenced by supply-side and demand-side determinants. The current primary healthcare capacity has been constrained by huge workload of limited health staff.

We recommend short-term strategies as follows.

First, increase patient awareness through empowerment and reduce time lag between screening and definitive diagnosis, thus reducing the gap between detected need, treatment initiation and crude coverage. Moreover, screening services should be extended from the current sub-district health centres to any public and private healthcare facilities for increased accessibility by all.

Second, minimize the gaps between crude and effective coverage through the application of Chronic Care Model, which requires a paradigm shift to team-based care, increased attention to non-pharmacological intervention and patient empowerment. Diversify screening services from sub-district health centres to any public and private healthcare facilities can increase the detected need indicator. And improve records on key outcome measurement, HbA1c and blood pressure.

Third, capitalize the potential opportunities from disruptive digital technology and innovations to support the current over-stretched health personnel.

In parallel, long-term strategies focusing on primary prevention to slow down the prevalence should be emphasized. These include:

First, strengthen campaign to modify lifestyle of diet calorie and total fat intake restrictions plus moderate-to-vigorous aerobic exercises can significantly improve diabetes risk factors; and that more intensive programs are more effective for higher risk group [49,50]. Thailand can capitalize the current high level of adequate physical activity in the population, in addition to other combined interventions.

Second, initiate and support population-based strategies which aim to achieve a small reduction in blood pressure in the entire population, by addressing risk factors, notably overweight, obesity, high sodium intake, low intake of potassium, unhealthy diet, high levels of alcohol consumption, low levels of physical activity [51,52]. In long run, these two recommendations will gradually slow down the increased prevalence of both conditions.

## Data Availability

Data cannot be shared publicly because of the confidential policy of the owner. Data
are available from the National Health Security Office, Thailand. Institutional Data
Access (contact via https://www.nhso.go.th) for researchers who meet the criteria for
access to confidential data.

## Acknowledgements

We would like to thank Waritta Wangbanjongkul, Anond Kulthanmanusorn, Shaheda Viriyathorn, Sataporn Julchoo, Mathudara Phaiyarom, Pigunkaew Sinam, Putthipanya Rueangsom, Theerawich Likitabhorn, Apipat Wiriya, and Parinda Seneerattanaprayul for their assistance in qualitative data collection, also, Jutatip Thungthnog, Poonchana Wareechai, and Pornpimol Sirimai for providing the data from National Health Security Office (NHSO) database. This publication has been supported by NHSO.

## Ethics

Ethics approval was granted by the Institute for the Development of Human Research Protection, Thailand. The ethics approval number is COA No. IHRP2019030, obtained on the 27th of March 2019.

## Patient and Public Involvement

It was not appropriate or possible to involve patients or the public in the design, or conduct, or reporting, or dissemination plans of our research.

