## Supplemental Table 1 for "Effective coverage of diabetes and hypertension in Thailand: challenges and recommendations"

**S1 Table. Number and profile of key informants, characteristics of study population, and disease prevalence**

**Number and profile of key informants**

| **Stakeholder groups** | **Male** | **Female** | **Total** |
| --- | --- | --- | --- |
| - Clinical specialist | 16 | 4 | 20 |
| - Healthcare providers and public health officers | 8 | 45 | 53 |
| - Academic | 2 | 5 | 7 |
| - Health insurance agencies | 3 | 2 | 5 |
| **Location of stakeholder’s affiliation** |  |  |  |
| - Central | 18 | 5 | 23 |
| - Regional | 11 | 51 | 62 |
| **Total** | 29 | 56 | 85 |
