## Supplemental Table 2 for "Effective coverage of diabetes and hypertension in Thailand: challenges and recommendations"

**S2 Table. Number of cases and effective coverage indicators for diabetes by sex and age group
2016-2019**

| **Indicators** | **Total** | **Sex** | | | **Age group** | | |
| --- | --- | --- | --- | --- | --- | --- | --- |
|  |  | **Female** | **Male** | **% Difference** | **< 60 years** | **≥ 60 years** | **% Difference** |
| **2016** | | | | | | | |
| Number of diabetes | 3,082,363 | 1,766,844 | 1,315,518 |  | 1,777,785 | 1,304,578 |  |
| Detected need | 2,077,382 (67.4%) | 1,394,114  (78.9%) | 679,874  (51.7%) | +27.2 | 925,245  (52.0%) | 1,148,743  (88.1%) | +36.1 |
| Crude coverage | 1,194,212 (38.7%) | 842,380  (47.7%) | 351092  (26.7%) | +21.0 | 507,869  (28.6%) | 685,603  (52.6%) | +24.0 |
| Effective coverage | 250,636 (8.1%) | 172,802  (9.8%) | 77836  (5.9%) | +3.9 | 84,833  (4.8%) | 165,805  (12.7%) | +7.9 |
| **2017** | | | | | | | |
| Number of diabetes | 3,131,096 | 1,797,448 | 1,333,648 |  | 1,778,317 | 1,352,779 |  |
| Detected need | 2,188,805 (69.9%) | 1,459,842  (81.2%) | 725,436  (54.4%) | +26.8 | 951,503  (53.5%) | 1,233,775  (91.2%) | +37.7 |
| Crude coverage | 1,348,416 (43.1%) | 944287  (52.5%) | 403331  (30.2%) | +22.3 | 562,464  (46.1%) | 785,154  (58.0%) | +11.9 |
| Effective coverage | 327,563 (10.5%) | 223363  (12.4%) | 104073  (7.8%) | +4.6 | 109,836  (6.2%) | 217,600  (16.1%) | +9.9 |
| **2018** | | | | | | | |
| Number of diabetes | 3,184,672 | 1,834,106 | 1,350,565 |  | 1,770,476 | 1,414,196 |  |
| Detected need | 2,290,339 (71.9%) | 1,518,332  (82.8%) | 768,533  (56.9%) | +25.9 | 971,610  54.9%) | 1,315,255  (93.0%) | +38.1 |
| Crude coverage | 1,436,386 (45.1%) | 1,000,436  (54.5%) | 435,187  (32.2%) | +22.3 | 581,638  32.9%) | 853,985  (60.4%) | +27.5 |
| Effective coverage | 375,892 (11.8%) | 254,619  (13.9%) | 121,149  (9.0%) | +4.9 | 122,979  (6.9%) | 252,789  (17.9%) | +11.0 |
| **2019** | | | | | | | |
| Number of diabetes | 3,215,033 | 1,847,911 | 1,367,122 |  | 1,754,109 | 1,460,923 |  |
| Detected need | 2,402,526 (74.7%) | 1,586,027  (85.8%) | 816,499  (59.7%) | +26.1 | 994,232  (56.7%) | 1,408,294  (96.4%) | +39.7 |
| Crude coverage | 1,601,761 (49.8%) | 1,109,419  (60.0%) | 492,342  (36.0%) | +24.0 | 631,358  (36.0%) | 970,403  (66.4%) | +30.4 |
| Effective coverage | 374,668 (11.7%) | 252,666  (13.7%) | 122,002  (8.9%) | +4.8 | 116,199  (6.6%) | 258,469  (17.7%) | +11.1 |
