## supplemental Table 3 for "Effective coverage of diabetes and hypertension in Thailand: challenges and recommendations"

**S3 Table. Number of cases and the effective coverage indicators for hypertension by sex and age group 2016-2019**

| **Indicators** | **Total** | **Sex** | | | **Age group** | | |
| --- | --- | --- | --- | --- | --- | --- | --- |
|  |  | **Female** | **Male** | **% Difference** | **< 60 years** | **≥ 60 years** | **% Difference** |
| **2016** | | | | | | | |
| Number of hypertension | 8,745,883 | 4,369,462 | 4,376,421 |  | 4,879,866 | 3,866,018 |  |
| Detected need | 4,273,732 (48.9%) | 2,725,539  (62.4%) | 1,541,727  (35.2%) | +27.2 | 1,670,596  (34.2%) | 2,596,670  (67.2%) | +33.0 |
| Crude coverage | 1,954,560 (22.3%) | 1,325,919  (30.3%) | 627,947  (14.3%) | +16 | 713,386  (14.6%) | 1,240,480  (32.1%) | +17.5 |
| Effective coverage | 984,414 (11.3%) | 681291  (15.6%) | 304344  (7.0%) | +8.6 | 368,982  (7.6%) | 616,653  (16.0%) | +8.4 |
| **2017** | | | | | | | |
| Number of hypertension | 8,892,742 | 4,465,895 | 4,426,847 |  | 4,881,586 | 4,011,156 |  |
| Detected need | 4,473,514 (50.3%) | 2,842,813  (63.7%) | 1,624,719  (36.7%) | +27 | 1,705,634  (34.9%) | 2,761,898  (68.9%) | +34.0 |
| Crude coverage | 2,200,512 (24.7%) | 1,484,092  (33.2%) | 715,686  (16.2%) | +17 | 783,038  (16.0%) | 1,416,740  (35.3%) | +19.3 |
| Effective coverage | 1,170,216 (13.2%) | 802,219  (18.0%) | 369,577  (8.3%) | +9.7 | 427,904  (8.8%) | 743,892  (18.5%) | +9.7 |
| **2018** | | | | | | | |
| Number of hypertension | 9,050,043 | 4,553,995 | 4,496,047 |  | 4,879,447 | 4,170,595 |  |
| Detected need | 4,683,544 (51.8%) | 2,962,358  (65.0%) | 1,715,246  (38.2%) | +26.8 | 1,739,461  (35.6%) | 2,938,143  (70.4%) | +34.8 |
| Crude coverage | 2,397,570 (26.5%) | 1,611,584  (35.4%) | 785,290  (17.5%) | +17.9 | 827,352  (17.0%) | 1,569,522  (37.6%) | +20.6 |
| Effective coverage | 1,362,928 (15.1%) | 927,054  (20.4%) | 433,050  (9.6%) | +10.8 | 481,538  (9.9%) | 878,566  (21.1%) | +11.2 |
| **2019** | | | | | | | |
| Number of hypertension | 9,161,148 | 4,634,311 | 4,526,837 |  | 4,816,575 | 4,344,572 |  |
| Detected need | 4,886,990 (53.3%) | 3,080,586  (66.5%) | 1,806,404  (39.9%) | +26.6 | 1,764,823  (36.6%) | 3,122,167  (71.9%) | +35.3 |
| Crude coverage | 2,678,785 (29.2%) | 1,791,927  (38.7%) | 886,858  (19.6%) | +19.1 | 895,393  (18.6%) | 1,783,392  (41.0%) | +22.4 |
| Effective coverage | 1,434,941 (15.7%) | 970,255  (20.9%) | 464,686  (10.3%) | +10.6 | 490,978  (10.2%) | 943,963  (21.7%) | +11.5 |
